# Radiologist-AI workflow can be modified to reduce the risk of medical malpractice claims

**DOI:** 10.1101/2025.06.14.25329278

**Authors:** Michael H. Bernstein, Brian Sheppard, Michael A. Bruno, Parker S. Lay, Grayson L. Baird

## Abstract

**Background:** Artificial Intelligence (AI) is rapidly changing the legal landscape of radiology. Results from a previous experiment suggested that providing AI error rates can reduce perceived radiologist culpability, as judged by mock jury members (4). The current study advances this work by examining whether the radiologist’s behavior also impacts perceptions of liability. Methods. Participants (n=282) read about a hypothetical malpractice case where a 50-year-old who visited the Emergency Department with acute neurological symptoms received a brain CT scan to determine if bleeding was present. An AI system was used by the radiologist who interpreted imaging. The AI system correctly flagged the case as abnormal. Nonetheless, the radiologist concluded no evidence of bleeding, and the blood-thinner t-PA was administered. Participants were randomly assigned to either a 1.) single-read condition, where the radiologist interpreted the CT once after seeing AI feedback, or 2.) a double-read condition, where the radiologist interpreted the CT twice, first without AI and then with AI feedback. Participants were then told the patient suffered irreversible brain damage due to the missed brain bleed, resulting in the patient (plaintiff) suing the radiologist (defendant). Participants indicated whether the radiologist met their duty of care to the patient (yes/no). Results. Hypothetical jurors were more likely to side with the plaintiff in the single-read condition (106/142, 74.7%) than in the double-read condition (74/140, 52.9%), p=0.0002. Conclusion. This suggests that the penalty for disagreeing with correct AI can be mitigated when images are interpreted twice, or at least if a radiologist gives an interpretation before AI is used.

## Introduction

Artificial Intelligence (AI) is rapidly changing the legal landscape of radiology [1,2]. One largely overlooked aspect of this change is how the human factors of AI implementation can impact legal liability. In one experiment, participants imagined viewing their mammogram report where a radiologist report disclosed a negative (BIRADS 1) interpretation. In some conditions, an AI report was additionally provided. Participants were asked whether they would consider consulting an attorney for a potential lawsuit if they subsequently learned they did have breast cancer. Participants were less likely to say they would pursue a consult if AI’s error rates were (versus were not) also included in the AI report (versus not) [3]. Results from another experiment suggested that providing AI error rates can reduce perceived radiologist culpability, as judged by mock jury members [4]. Taken together, these studies suggest that information about AI’s performance modulate legal liability. The current study advances this work by examining whether the radiologist’s behavior also impacts perceptions of liability.

## Methods

Participants (n=282, 156 (55.7% female)) recruited online (Prolific.com) read about a hypothetical case where a 50-year-old who visited the Emergency Department with acute neurological symptoms received a brain CT scan to determine if bleeding was present. An AI system was used by the radiologist who interpreted imaging. The AI system correctly flagged the case as abnormal. Nonetheless, the radiologist concluded no evidence of bleeding, and the blood-thinner t-PA was administered.

Participants were randomly assigned to either a single-read condition where the radiologist interpreted the CT once after seeing AI feedback; or a double-read condition where the radiologist interpreted the CT twice, first without AI and then with AI feedback.

Participants were then told the patient suffered irreversible brain damage due to the missed brain bleed, resulting in the patient (plaintiff) suing the radiologist (defendant). Participants indicated whether the radiologist met their duty of care to the patient (yes/no), consistent with typical questions regarding breach of duty of care that appear on jury verdict forms in medical malpractice trials. Ethics committee/IRB of Brown University Health waived ethical approval for this work. All data produced in the present study are available upon reasonable request to the authors. More detail is provided elsewhere [4].

## Results

Illustrated in Figure 1, hypothetical jurors were more likely to side with the plaintiff in the single-read condition (106/142, 74.7%, (95% CI [66.8, 88.1]) compared to the double-read condition (74/140, 52.9%, (95% CI [44.6, 61.0]), p=0.0002.

**Figure 1.**
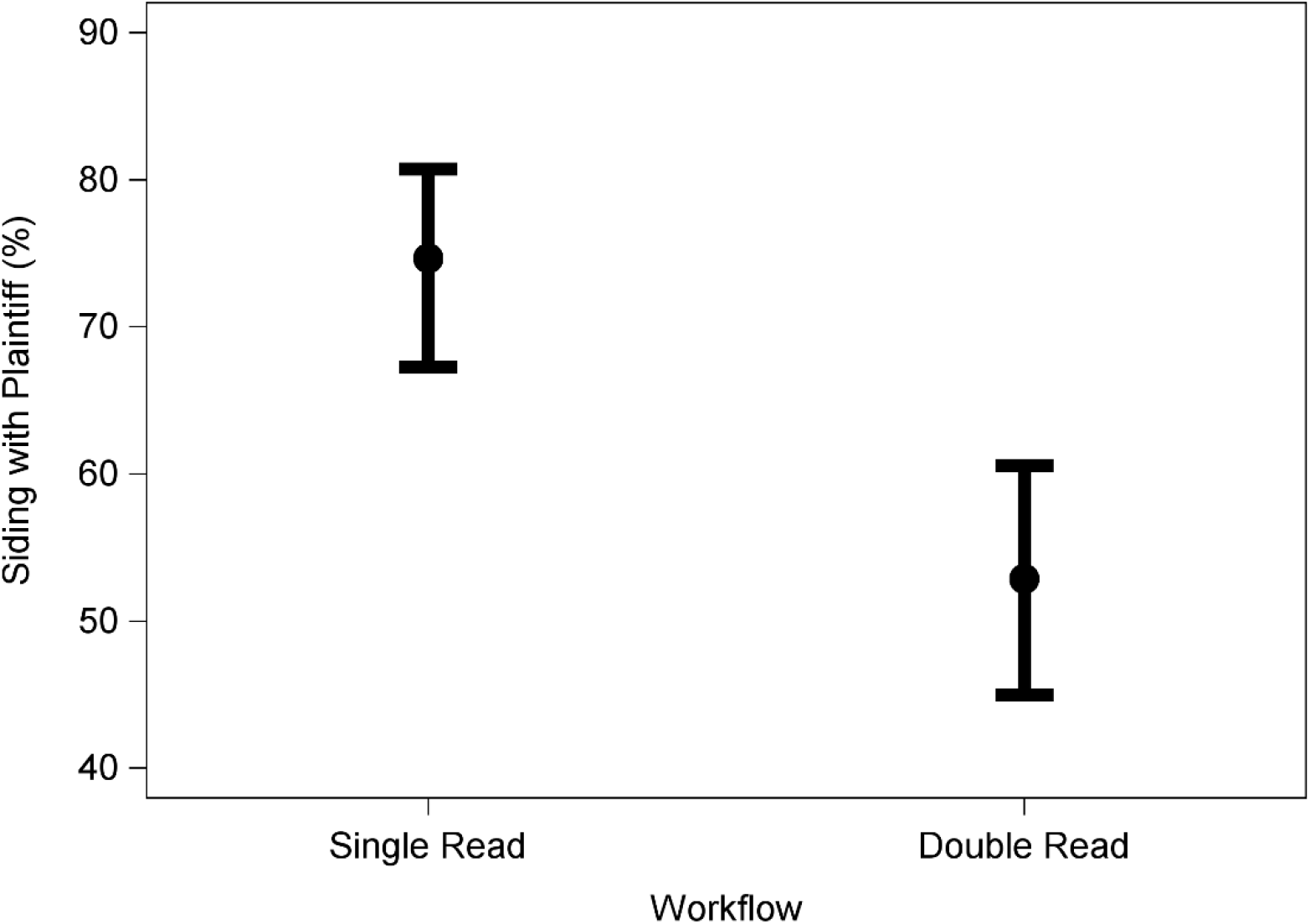
Siding with the plaintiff between Single and Double reading Workflows. Note. Bars represent upper and lower bounds of 95% confidence intervals.

## Discussion

Mock jurors were more likely to believe that the radiologist met their duty of care when a false negative interpretation occurred after reading an image twice–once without AI and then once with AI–relative to only reading the image once with AI. The magnitude of this effect was large (74.7% vs. 52.9%). Moreover, participants sided with the plaintiff in the double-read condition (52.9%) almost as often as if no AI had been used (56.3%) (see [4]). This suggests that the penalty for disagreeing with correct AI can be mitigated when images are interpreted twice, or at least if a radiologist gives an interpretation before AI is used.

The generalizability of these findings to medical malpractice cases in actual jury trials is unknown. It is also unknown if the double-read condition will reduce liability when AI also fails to detect an abnormality. Finally, whether the double-read condition reduced legal risk because the images were read twice or because of the order in which the two reads occurred (AI first or radiologist first) is unknown. Nonetheless, these findings could have important real-world implications. Jurors are not ordinarily allowed to learn about a doctor’s performance on prior cases [5], but they can learn about their performance on the patient’s case.

AI invites challenging questions regarding medical malpractice among radiologists [1].

## Data Availability

All data produced in the present study are available upon reasonable request to the authors.

